# Predictive Performance Precision Analysis in Medicine: Identification of low-confidence predictions at patient and profile levels (MED3pa I)

**DOI:** 10.1101/2025.08.22.25334254

**Authors:** Olivier Lefebvre, Félix Camirand Lemyre, Jean-François Ethier, Lyna Hiba Chikouche, Ludmila Amriou, Dan Poenaru, Martin Vallières

## Abstract

**Objective:** Artificial Intelligence models are increasingly used in healthcare, yet global performance metrics can mask variations in reliability across individual patients or subgroups with shared attributes, called **patient profiles**. This study introduces MED3pa, a method that identifies when models are less reliable, allowing clinicians to better assess model limitations.

**Materials and Methods:** We propose a framework that estimates predictive confidence using three combined approaches: Individualized (IPC), Aggregated (APC), and Mixed Predictive Confidence (MPC). IPC estimates confidence for each patient, APC assesses it across profiles, and MPC combines both. We evaluate our method on four datasets: one simulated, two public, and one private clinical dataset. Metrics by Declaration Rate (MDR) curves show how performance changes when retaining only the most confident predictions, while interpretable decision trees reveal profiles with higher or lower model confidence.

**Results:** We demonstrate our method in internal, temporal, and external validation settings, as well as through a clinical example. In internal validation, limiting predictions to the 93% most confident cases improved sensitivity by 14.3% and the AUC by 5.1%. In the clinical example, MED3pa identified a patient profile with high misclassification risk, demonstrating its potential for safer deployment.

**Discussion:** By identifying low-confidence predictions, our framework improves model reliability in clinical settings. It can be integrated into decision support systems to help clinicians make more informed decisions. Confidence thresholds help balance model performance with the proportion of patients for whom predictions are considered reliable.

**Conclusion:** Better leveraging confidence in model predictions could improve reliability and trustworthiness, supporting safer and more effective use in healthcare.

## 1 BACKGROUND AND SIGNIFICANCE

Artificial intelligence (AI) has the potential to transform healthcare, with clinical models playing a key role in advancing precision medicine by helping diagnosis and prognosis^1–3^. These models typically undergo internal training and validation, followed by external validation across diverse datasets^1,4–6^. However, as highlighted by multiple studies^7–11^, this standard pipeline does not guarantee generalizability due to variations in populations and contexts. Machine learning (ML) methods often assume congruence between training and application distributions; this assumption, however, is not always met in real-world applications. This discord is often attributed to dataset shifting, where the joint distribution of inputs and outputs differs between training and testing phases^12^. Another deficiency in AI models is their focus on estimating local averages by mapping the predictor space onto a latent dimension where their relationship with the desired outcome is accentuated. Although this approach optimizes global performance metrics, it may overlook underrepresented subpopulations and fail to account for individual variability^13^. As a result, predictions may be unreliable for some patients, meaning that the model’s output is less likely to accurately reflect the patient’s true outcome. Moreover, predictive performance may vary between patients even if the model is well-calibrated. Some **patient profiles** — defined as subgroups of patients with shared features such as age or comorbidities — may systematically exhibit higher prediction errors. Although patient profiles are defined by rule-based patient characteristics, model performance is used to identify which profiles are associated with systematically higher or lower predictive performance. It then becomes imperative to conduct a comprehensive analysis to identify and characterize such low-confidence predictions. Here, confidence reflects the expected reliability of the model’s output for each patient, with lower confidence indicating a higher risk of incorrect prediction. This analysis should be performed prior to model deployment and maintained continuously as clinical contexts and data distributions evolve.

Multiple factors can cause a model to make erroneous predictions. Even when deployment data follow the same distribution as the training data, some cases lack discriminative information, and the inherent variability between outcomes and predictors cannot be reduded, which is known as aleatoric uncertainty. Epistemic uncertainty can also lead to incorrect prediction, arising from limitations within the model itself, including misspecification or insufficient representation of certain patient subgroups. As a result, the model may be correctly specified for the dominant subpopulation but misspecified for varying underrepresented subgroups or individuals. Even with correct model specification, predictions may remain uncertain for certain patient profiles, e.g., logistic regression when predicted probabilities are near 0.5.

Figure 1 illustrates how these sources of uncertainty can manifest in practice. Figure 1a shows examples within the training data where the model 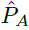 is less accurate due to limited discriminative information. Figure 1b highlights how temporal or external datasets can diverge from the training distribution, demonstrating the importance of contextual validation. Even if the model 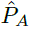 is correctly specified for the training distribution (Set A), its predictions are less reliable for patients with higher values of **x**_1_ (red rectangle) due to subgroup underepresentation. Temporal datasets refer to future data collected from the same source, while external datasets originate from different institutions or populations. Models developed in a specific context may degrade or fail to generalize, as seen in *Vela et al.*^14^, who observed temporal performance declines even in stable environments, reinforcing the need for continuous monitoring.

**Figure 1:**
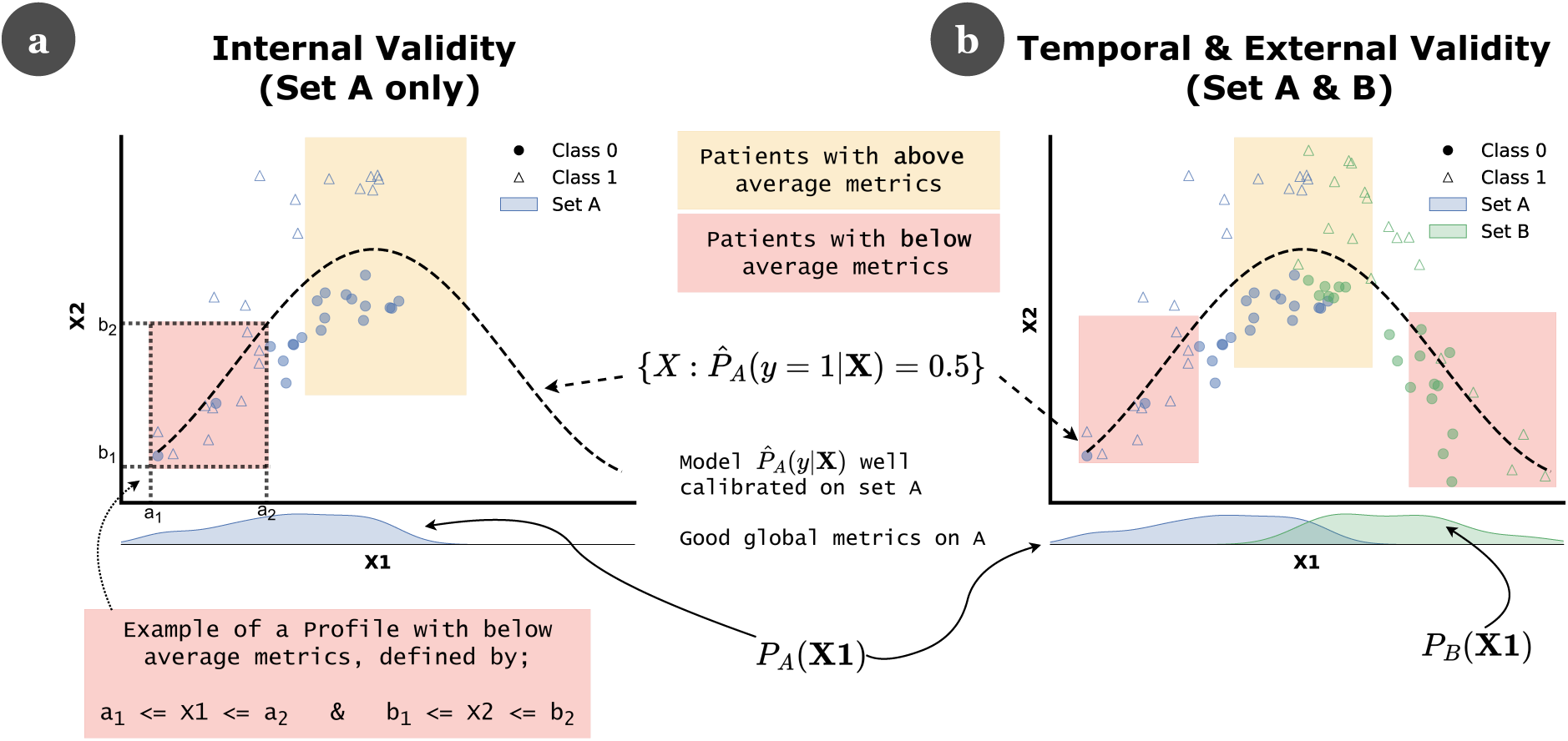
Illustration of model reliability across Internal and Temporal/External contexts. (a) Internal validity: Scatter plot showing a classification model trained on two features (X1 and X2) to separate blue triangles and circles. The dashed curve represents the discriminant boundary of the model, defined by 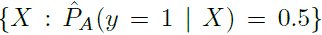, where 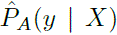 is trained and calibrated on Set A. In the central region (yellow rectangle), classes are well separated and the model is more reliable. On the left (red rectangle), class overlap leads to lower model reliability. (b) Temporal & External validity: Additional data points from Set B (green) reveal a new region on the right (red rectangle) where the model becomes unreliable, due to an overlap between the two classes not present in the original data. The marginal distributions *P_A_*(*X*1) and *P_B_*(*X*1) illustrate a shift in feature *X*1 between Sets A and B, contributing to changes in performance across profiles.

Several approaches have been proposed to improve predictive model reliability and interpretability, particularly under of distribution shifts and performance disparities. *Ginsberg et al.*^12^ introduced *Detectron*, a hypothesis test detecting harmful covariate shifts by analyzing an ensemble of constrained disagreement classifiers. *Shashikumar et al.*^15^ proposed *COMPOSER*, a sepsis model using conformal prediction to flag unfamiliar cases as indeterminate, improving model deployment across diverse populations. Conformal prediction provides regions containing the true outcome with a specified confidence level^16^. Recent works extended conformal prediction to handle covariate shifting, enabling confident predictions even when testing data differ from training^17–19^. Conformal prediction provides guarantees on predictions by constructing calibrated prediction regions from past data, but it focuses on producing valid prediction sets rather than characterizing specific patients associated with higher uncertainty.

In contrast, uncertainty quantification (UQ) techniques are specifically designed to estimate predictive uncertainty, aiming to provide reliable predictions^20–24^. These methods typically estimate epistemic and/or aleatoric uncertainty, often during model training, and rely on assumptions on the underlying learning algorithm. As a result, they are not model-agnostic and primarily characterize uncertainty within the training data distribution. Furthermore, those uncertainty estimates do not necessarily reflect whether a trained model will produce an incorrect prediction on a given test instance^25–27^, and the quality of uncertainty estimation degrades under distribution shifts^28,29^.

An alternative line of work reframes uncertainty estimation as a prediction task, aiming to forecast if a model’s prediction is likely to be erroneous on new inputs. These methods focus on predicting model performance rather than uncertainty intrinsic to the model or data. Most approaches formulate this task as a binary classification problem, predicting whether a model’s output is correct for a given instance^25,30–34^. Framing error prediction as a binary outcome however ignores the magnitude of prediction errors. Using continuous error measures can instead emphasize highly erroneous high probability predictions, assigning lower confidence to predictions that the initial model makes with high confidence but are likely to be wrong.

The effectiveness of these methods is often evaluated using accuracy-rejection curves^30,33,35,36^, which measures the accuracy of a base classifier after progressively rejecting predictions with lowest confidence. Accuracy-based evaluation may however be misleading in certain settings, such as highly imbalanced datasets. Alternative rejection curves have been proposed based on other performance metrics, such as precision, recall or AUROC^31,37,38^.

Complementary to UQ, the hidden stratification literature investigates performance disparities across subgroups, allowing a more interpretable explanation of model limitations. Some approaches rely on manually defined subgroups^39,40^, which may restrain the efficiency of subgroups discovery. Several automatic methods have been proposed without requiring predefined rules, however many of these are based on clustering representations in the latent space of neural networks^41–45^, restricting their applicability to neural network-based models. To address this limitation, QUEST^46^ was proposed as a model-agnostic hidden stratification method that uses decision tree induction to identify easily interpretable subgroups associated with discrete levels of uncertainty. Discretizing uncertainty into a small number of bins may limit the granularity of uncertainty estimates, potentially resulting in less precise subgroup characterization.

In this work, we introduce the *Predictive Performance Precision Analysis in Medicine* (MED3pa) method. MED3pa is a model-agnostic framework for the post-hoc identification and characterization of potential failures of a preexisting predictive model, referred to as the **BaseModel**, which represents a well-trained and calibrated binary classification model. MED3pa identifies patient profiles for which the BaseModel exhibits reduced performance, supporting both pre-deployment evaluation and post-deployment monitoring. This article presents the first phase of MED3pa, focusing on identifying when and for whom a BaseModel is less reliable by detecting low-confidence predictions and revealing the most affected patient profiles. Future work will explore the underlying causes of these reliability gaps and explore strategies to improve reliability across patient profiles.

## 2 OBJECTIVE

### The overarching goal of MED3pa is not to exclude patients from care, but to improve the inclusivity and reliability of model predictions for *all* patients

A prediction may have a high probability of being incorrect not necessarily because the model is unreliable, but due to inherent uncertainty or variability in the data such as underrepresentation of certain profiles or high outcome variability. By identifying patients for whom a model may be less reliable, we aim to better understand and address its limitations. However, in this first work, we focus on identifying these patients and, for now, withholding predictions for them to explore how excluding uncertain cases can balance performance improvements with patient inclusion. Specifically, we aim to:

- **develop a method** to detect and characterize patient profiles associated with a high likelihood of inaccurate predictions, to provide insights on prediction reliability in clinical models;
- **demonstrate the method** on a simulated dataset, to validate its functionality in a controlled setting; and
- **apply the method** to real clinical datasets, to assess its effectiveness in a clinical context.

#### Intended uses

The MED3pa method offers a range of applications aimed at improving our understanding and utilization of a pre-existing BaseModel, both for initial validation and continuous monitoring. While not exhaustive, key applications include:

1. **assessing applicability and reliability**: evaluate if the BaseModel is suitable for specific clinical contexts;
2. **investigating patient profiles**: identify and analyze patient profiles for which the BaseModel demonstrates reduced performance; and
3. **daily clinical application**: evaluate whether the BaseModel can be used for a given patient, ensuring predictive capabilities align with patient care objectives.

## 3 MATERIALS AND METHODS

### 3.1 Predictive Performance Precision Analysis

We consider a generic binary classification model that has been previously trained, calibrated, and validated, denoted as the **BaseModel**. Our goal is to evaluate one such model on new, unseen data from the deployment context, referred to as **evaluation** sets. These sets may also include additional patient characteristics, such as sociodemographics, which can later be used to explore hypotheses about performance disparities. While global metrics such as the Area Under the Receiver Operating Characteristic curve (AUC), sensitivity, and specificity are typically used to assess performance in new contexts, they may overlook patient-level variability. To address this, we propose an approach assessing predictive confidence at both individual patient and customizable patient profile levels within these evaluation sets, identifying where the model is less reliable. This allows exploration of performance improvements by selectively withholding predictions, while balancing metric gains and patient inclusion for which the BaseModel can be more safely applied.

To illustrate our method, we first use a simulated dataset to visualize key concepts (data generation process is described in Supplementary Material A). Figure 2a shows the generated training/testing data and the classification regions produced by the BaseModel, which is a random forest classifier.

**Figure 2:**
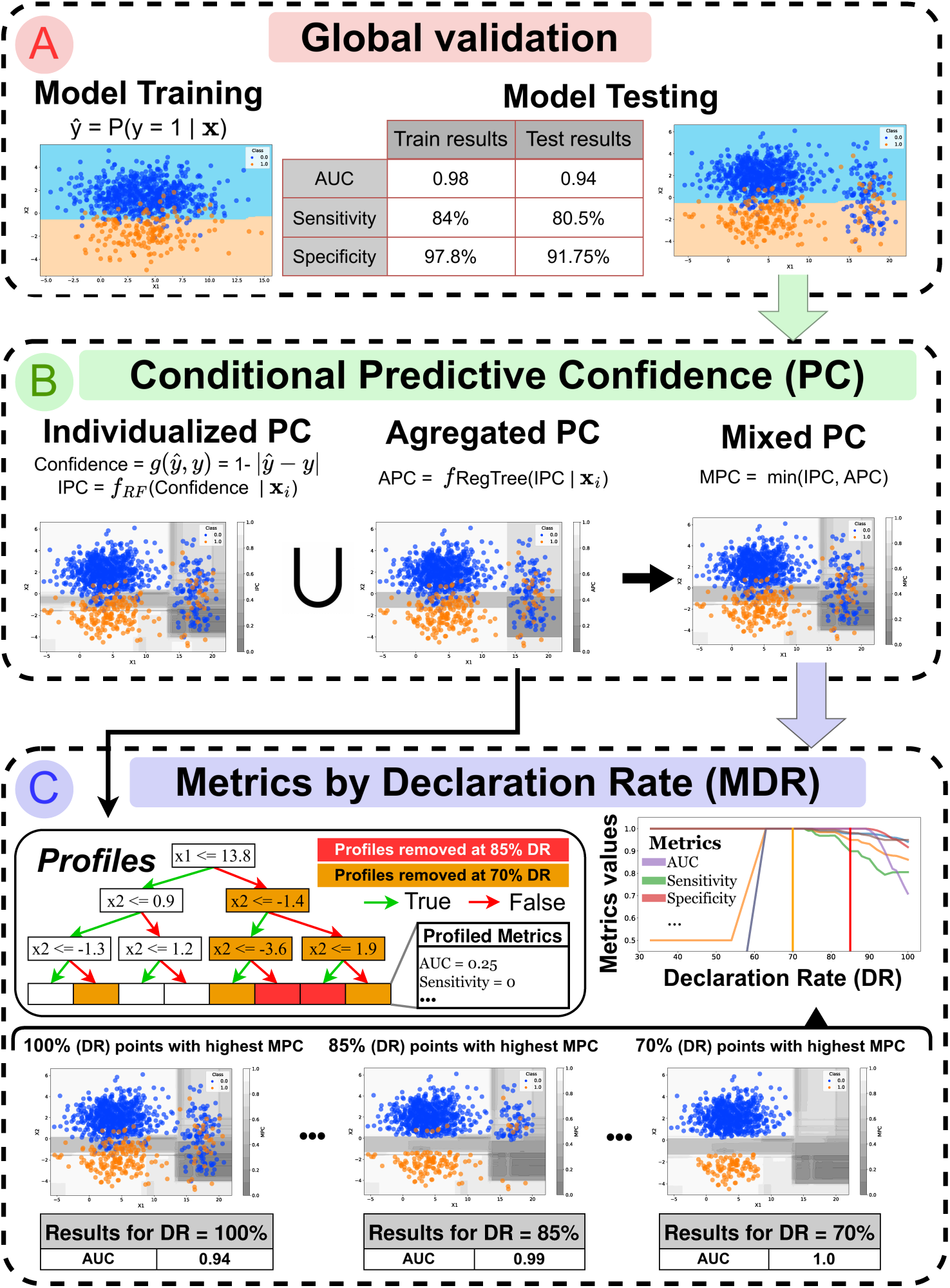
Overview of the proposed MED3pa method for identifying and managing predictive confidence using a simulated dataset. (a) Conventional global validation: A BaseModel is trained on a training dataset and evaluated on a separate test dataset, with performance reported as global metrics. (b) Conditional Predictive Confidence: Example confidence estimates on the test set using the Individualized Predictive Confidence (IPC), Aggregated Predictive Confidence (APC), and Mixed Predictive Confidence (MPC) models, each highlighting different aspects of predictive confidence. (c) Metrics by Declaration Rate (MDR): MDR curves visualize how performance metrics evolve as increasingly uncertain predictions (based on the MPC) are removed. The APC model is also used to identify interpretable patient profiles associated with low confidence. The bottom panel illustrates three examples of DR on the testing data, with DR = 100% where no patient is excluded, and DR = 70% where 30% of patients with lowest confidence are excluded from the use of the BaseModel.

#### 3.1.1 Defining confidence

We define the predictive confidence of a BaseModel for patient i as 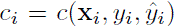, where **x***_i_* denotes the patient’s characteristics, *y_i_* the true outcome, and 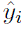 the prediction generated by the BaseModel. This confidence metric quantifies the predictive performance of the BaseModel given patient characteristics. This confidence metric can be binary (e.g. 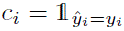) or continuous (e.g. 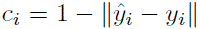), where 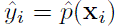 is the BaseModel’s prediction and *y_i_* is the true label. It may also be the predicted probability itself when the true outcome is unknown. However, our objective is to predict systematic model failures across patients, whereas the predicted probability primarily reflects the BaseModel scoring function and is less effective for failure prediction^20,25^. Empirical results supporting this observation are presented in Supplementary Material B. Asymmetrical metrics can also be used to reflect classification thresholds or misclassification costs. The choice of confidence measure depends on the deployment context.

#### 3.1.2 Individualized Predictive Confidence (IPC) Model

We then construct the **Individualized Predictive Confidence** (**IPC**) model, which predicts an estimate 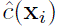 of the confidence metric for each new patient using features **x***_i_*. To train IPC, we first apply the BaseModel on a subset of the evaluation set and compute for each patient *i* a confidence value *c*(*x_i_*) using our chosen confidence metric. The IPC model is then trained to predict the confidence using patient features. IPC can be any classification model if the confidence metric is discrete, or a regression model if continuous.It is advisable to choose a model with similar or less capacity than the BaseModel to avoid overfitting confidence predictions. At deployment, IPC tales only features *x_i_* to provide an estimate of the confidence value for each patient before applying the BaseModel.

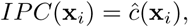

where 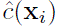 is the predicted confidence. Figure 2b shows an example using a random forest regressor as IPC to predict a confidence score defined as 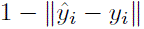.

#### 3.1.3 Aggregated Predictive Confidence (APC) Model

Although the IPC model provides confidence estimates for individual patients, it does not provide any mechanism to identify groups of patients with similar confidence patterns. Identifying low-confidence profiles highlights disadvantaged profiles and helps investigate the sources of these discrepancies. To address this, we introduce the **Aggregated Predictive Confidence** (**APC**) model. The APC model builds confidence profiles using supervised learning, where the target is the confidence values defined by the IPC model. A CART model (classification or regression depending on the confidence metric) is trained on the same subset of the evaluation set as the IPC, partitioning patients into profiles (represented by the leafs of the tree) that share similar expected confidence levels. For any patient profile *S_i_*, APC outputs the average IPC-predicted confidence within that profile:

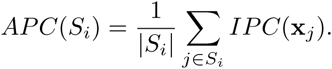

For an individual patient, APC outputs the average confidence associated with the smallest profile containing that patient, corresponding to a leaf leaf of the induced tree. The generation of these confidence profiles follows a rule-based, tree-structured approach similar to *QUEST*^46^. In APC, each profile is however associated with an overall confidence value, which is used to rank profiles by confidence rather than discretizing uncertainty into categories such as high versus low. Tree-based models are particularly valuable for producing interpretable profiles and computing APC values. Profile granularity is adjusted by changing tree depth and should be chosen contextually, reflecting the desired level of detail. Similar to QUEST^46^, it can also be determined by identifying the optimal trade-off between the number of profiles and the accuracy of predicted confidence. APC profiles are constructed using the patient characteristics in the evaluation set, which may include the features used by the BaseModel as well as additional patient variables. Figure 2b illustrates a regression tree-based APC, while Figure 2c shows the resulting profiles.

#### 3.1.4 Mixed Predictive Confidence (MPC) Model

The IPC model provides patient-specific confidence estimates, but these can vary widely between patients. In contrast, the APC model offers more consistent estimates by grouping similar patients into profiles, although it may miss important individual differences. By combining both models, we can achieve a better balance between individual precision and overall stability in estimating prediction reliability for each patient. We propose the **Mixed Predictive Confidence** (**MPC**) model, defined as:

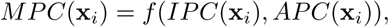

where *f* (*IPC*(**x***_i_*)*, APC*(**x***_i_*)) combines IPC and APC predictions. A simple approach is taking the minimum confidence, but other methods such as averaging could be used depending on context. MPC balances individual estimates with broader profile-level trends. Figure 2b illustrates an MPC model using the minimum strategy, highlighting both low-confidence profiles and individual points.

Training procedures for **IPC**, **APC** and **MPC** depend on intended use, whether to capture individual, profiled, or combined confidence. Further details are provided in Supplementary Material C.

#### 3.1.5 Metrics by Declaration Rate (MDR)

Once confidence estimates are available and low-confidence patients are identified in the evaluation subset, it remains necessary to determine minimum acceptable confidence levels before deploying the BaseModel. To guide this, we introduce **Metrics by Declaration Rate** (**MDR**), a generalization of the *Accuracy-Rejection Curves*^30,35^. MDR uses the remaining portion of the evaluation set not used for confidence model training, and assesses predictive performance by gradually excluding the least confident predictions. The **Declaration Rate (DR)** refers to the proportion of patients retained. For each DR, we compute metrics (e.g., AUC, accuracy). For example, at DR = 60%, 40% of patients with lowest predicted confidence are excluded, and the BaseModel is evaluated on the remaining 60%.

To construct MDR curves, patients are ranked by predicted confidence (via IPC, APC, or MPC). For each DR ∈ [0, 1], patients with lowest confidence are incrementally removed, and metrics such as AUC, sensitivity, and specificity are computed in the remaining subset. Plotting metrics against DR yields MDR curves, visualizing how excluding the least confident predictions affects performance.

Ideally, well-calibrated confidence estimates produce monotonically increasing MDR curves as erroneous predictions are excluded. In practice, confidence models often identify some, but not all, uncertain predictions. MDR curves may then increase, peak, and decline as further exclusion offers no benefit or harms performance. The optimal DR depends on the objective and should balance performance gains and patient inclusion.

Figure 2c shows MDR curves increasing up to DR = 60%, then decrease. For example, if the goal is to increase AUC, DR = 90% may be optimal as it marks where performance plateaus. It also illustrates how different DR values translate into removing the least confident predictions from the evaluation set, where predictive confidence is defined using the Mixed Predictive Confidence (MPC). It computed as the minimum between the Individualized Predictive Confidence (IPC) and the Aggregated Predictive Confidence (APC), thereby enabling a trade-off between individual-level and profile-level confidence. At DR = 100%, the scatter plot shows all data points, with the background color indicating predictive confidence (dark gray being the least confident and white the most confident). At DR = 85%, the 15% least confident predictions are removed, including the two patient profiles highlighted in red. At DR = 70%, the 30% least confident are removed, including profiles highlighted in red and orange.

### 3.2 Experimental setup

We applied our method to two classification problems: In-Hospital Mortality (IHM) using public datasets, and One-Year Mortality (OYM) using a private dataset. Further experimental details and algorithmic components are provided in Supplementary Material D.

#### 3.2.1 In-Hospital Mortality Prediction

The first task predicts IHM for ICU patients 24 hours after admission, using 17 predictors from the Simplified Acute Physiology Score (SAPS) II^47^. We used MIMIC-IV^48^ and eICU-CRD^49^, two public datasets from U.S. hospitals. MIMIC-IV contains electronic health records (EHRs) from Beth Israel Deaconess Medical Center (2008-2019), while eICU-CRD includes 208 hospitals (2014-2015). After applying preprocessing and exclusion criteria following *Zeng et al.*^50^, the final MIMIC-IV dataset included 16,271 admissions. In eICU-CRD, hospitals with less than 225 admissions were excluded to ensure sufficient sample size, resulting in 2,583 admissions across 9 hospitals.

The IHM BaseModel is an XGBoost classifier trained on half of the 2008-2013 MIMIC-IV cohort (n = 4,476; 16.8% positive rate). It was calibrated, with the classification threshold selected by maximizing Youden’s J-index^51^. The confidence metric used is a sigmoidal function (Supplementary Material E). MPC was defined as the minimum confidence between IPC (random forest regressor) and APC (regression tree). The BaseModel was applied to the remaining 2008-2013 patients (n = 4,476; 17.4% positive rate) for **internal validation**, the 2014-2019 cohort (n = 7,319; 18.8% positive rate) for **temporal validation**, and to each eICU hospital (n = 2,583; 18.1% positive rate) for **external validation**. Further details on data processing, variable transformations, exclusion criteria, evaluation sets construction, and descriptive statistics are provided in Supplementary Material F.

#### 3.2.2 One-Year Mortality Prediction

The second task predicts OYM risk upon admission, using data from *Laribi et al.*^52^, comprising EHRs and administrative data from an integrated university hospital network in Sherbrooke, Quebec, Canada. We used the “AdmDemoDx” dataset, which includes 2 demographic variables, 10 admission characteristics, 85 comorbidities, and 147 admission diagnoses^52^.

Following *Laribi et al.*^52^, the BaseModel was trained on 2011-2017 admissions (n = 148,587; 13.9% positive rate) and validated on 2017-2021 admissions (n = 61,310; 12.5% positive rate). The BaseModel is a calibrated random forest classifier, with threshold optimization via Youden’s index^51^. The same confidence metric as IHM was applied, with random forest regressor for IPC, regression tree for APC, and minimum confidence strategy for MPC. Additional dataset details and descriptive statistics are provided in Supplementary Material G.

## 4 RESULTS

All methods presented in this study are implemented in the public Python package MED3pa, which provides tools for estimating predictive confidence, evaluating model performance using MDR curves, and identifying error-prone patient profiles. MED3pa is available at: https://pypi.org/project/MED3pa/. The package includes a simple example based on synthetic data^53^ generated from the *Laribi et al.* dataset. Additionally, the code used to generate all results and figures in this study is available at: https://github.com/MEDomics-UdeS/study_3pa. We evaluated the performance of MED3pa across all experimental settings considered in this study, assessing its ability to detect BaseModel prediction errors using AUC. Detailed results are provided in Supplementary Material B.

### 4.1 Demonstration of the method via In-Hospital Mortality Prediction

We applied MED3pa to a BaseModel developed to predict in-hospital mortality using MIMIC-IV^48^ and eICU-CRD^49^ datasets. First, we assessed **internal validity** on a MIMIC-IV subset sampled from the same distribution as training data. Next, we evaluated **temporal validity** using data from a later period, and finally tested **external validity** on the eICU-CRD dataset. While MDR curves can include any metric, we focused on five to simplify analysis and visualization. Accuracy is not shown in these curves, but as detailed in Supplementary Material H, it improved consistently across scenarios. We therefore prioritized other metrics to better capture performance nuances.

#### 4.1.1 Internal Validation

Internal validation assesses whether a model is valid within the same context as training data. As shown in the MDR curves in Figure 3, removing patients with lowest predicted confidence led to improvements in all performance metrics except **Positive Predictive Value (PPV)**. **Specificity** remained stable for DR above 93% with a relative increase of 1.2% compared to DR=100% (Δ*_rel_*% = +1.2%), then improved, plateauing near 100% at *DR = 30%*. **Negative Predictive Value (NPV)** behaved similarly, mostly increasing for DRs above 93% (Δ*_rel_*% = +4.4%). However, the PPV continually decreased as DR was reduced, suggesting that maximizing PPV favors a DR of 100%.

**Figure 3:**
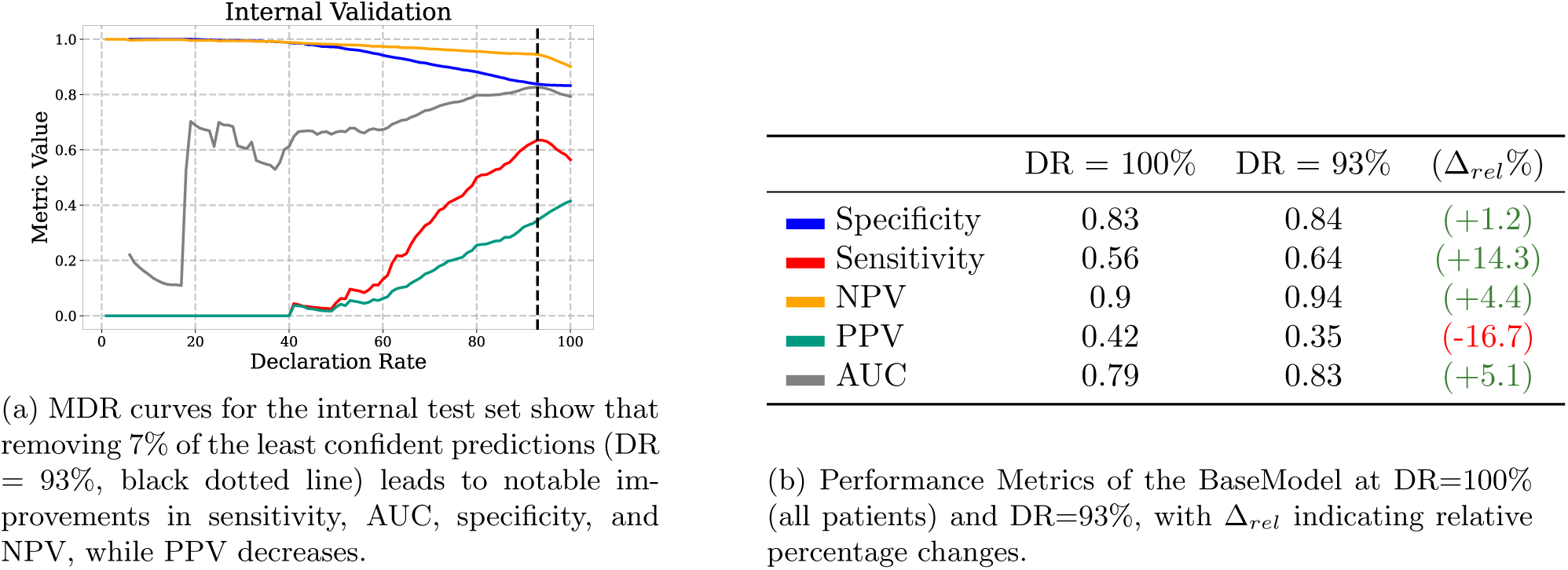
Internal validation of the BaseModel.

**Sensitivity** and **AUC** peaked at DR = 93%, with relative improvements of +14.3% and +5.1% respectively, before declining at lower DRs. Excluding the 7% least confident predictions would enhance all metrics except PPV, while more aggressive filtering would benefit only specificity and NPV.

#### 4.1.2 Temporal Validation

After deployment, model performance can degrade due to factors such as changes in patient populations or clinical workflows. Temporal validation ensures the model remains reliable as hospital characteristics evolve. As shown in Figure 4, specificity and NPV improved consistently by removing low-confidence patients, while other metrics tended to decline. For example, if the clinical objective is to maintain the same specificity as internal validation 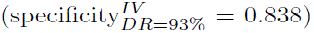, this would be achieved with a DR of 91%. However, for other objectives, retaining all patients may be preferable.

**Figure 4:**
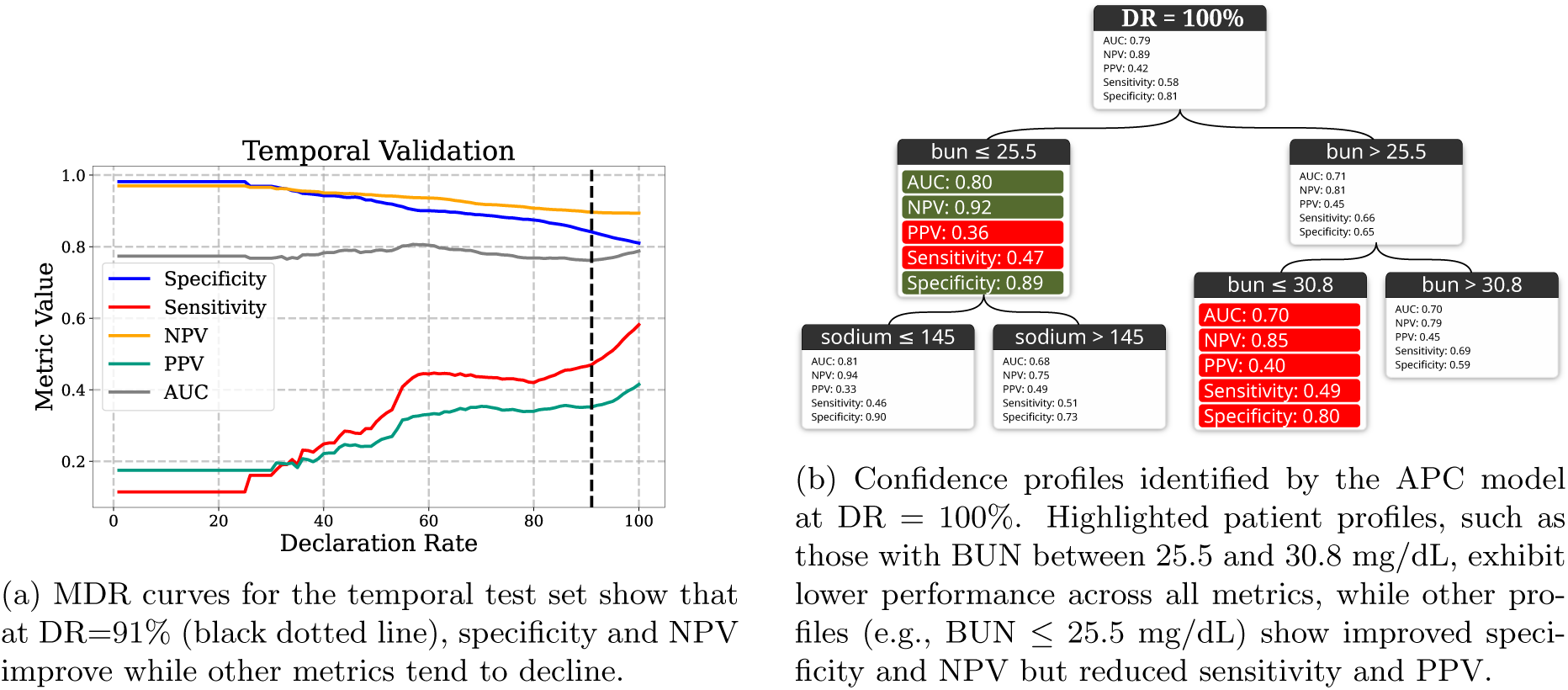
Temporal validation of the BaseModel.

Even when predicted confidences do not improve most metrics, confidence profiles still provide insights into model limitations and improvement targets. In Figure 4b), patients with a BUN between 25.5mg/dL-30.8mg/dL showed decreased performance across all metrics. Those with BUN ≤ 25.5 saw improved AUC, specificity, and NPV, but reduced sensitivity and PPV. This suggests the BaseModel is more confident in negative predictions for this profile, at the cost of positive prediction performance. Analyzing confidence profiles can support various goals depending on the context.

#### 4.1.3 External Validation

External validation simulates deploying the BaseModel in a context different from training data. Figure 5 shows MDR curves for three hospitals from the eICU-CRD database^49^ demonstrating metric improvements. For each hospital, we proposed an optimal DR and summarized metric changes compared to DR = 100% in Table 1.

**Figure 5:**
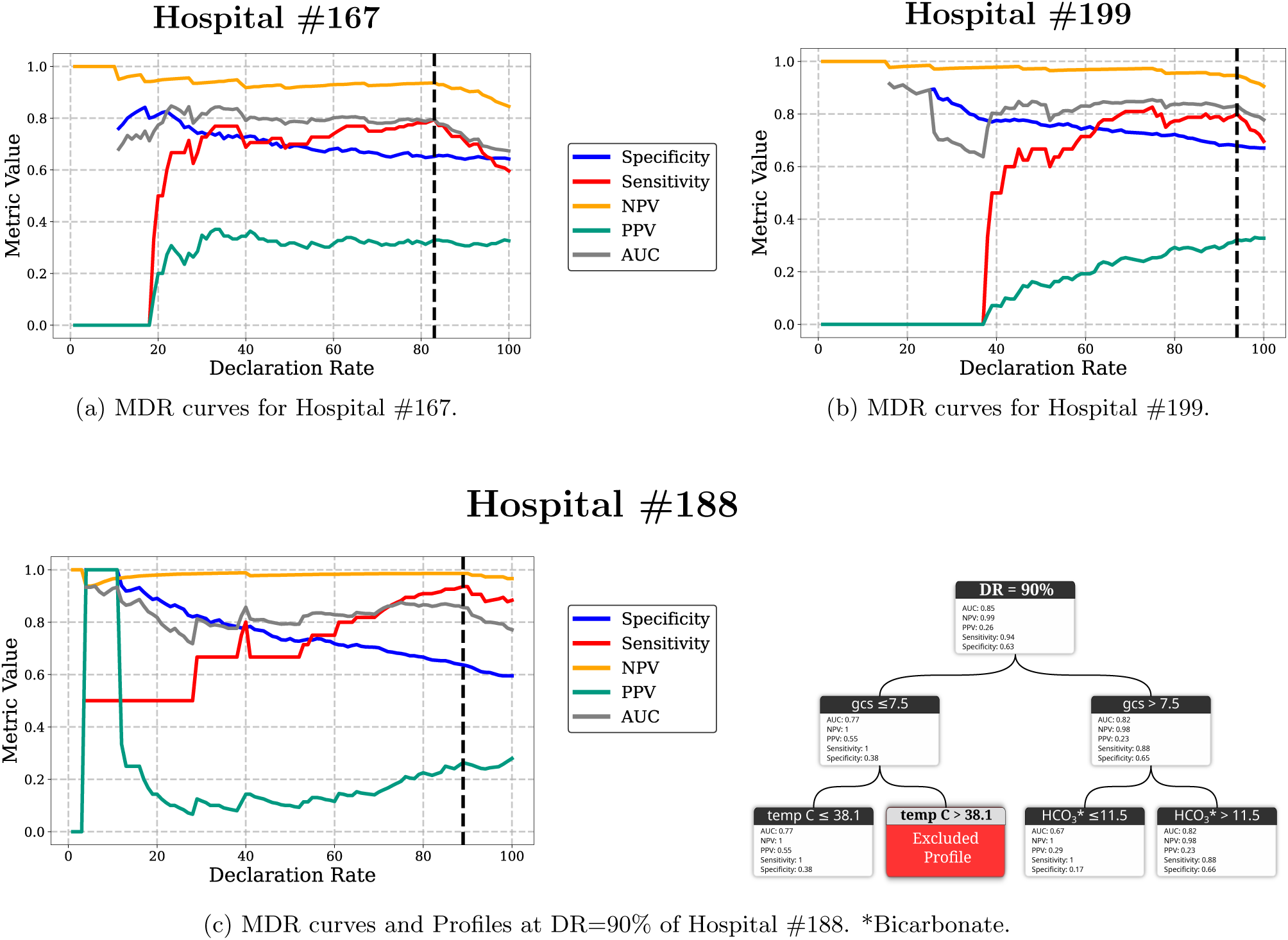
External validation of the BaseModel across three hospitals. (a-c) MDR curves for Hospitals #167, #199, and #188 show how model performance changes as the least confident predictions are excluded using MPC-based predictions. Most metrics demonstrate improvements up to a certain DR beyond which gains plateau or decline, except for PPV, which tends to decrease.

**Table 1:** External validation results across three eICU-CRD hospitals. For each hospital, we report model performance at DR = 100% (all patients included) and at the suggested DR, as well as the relative percentage change (Δ*_rel_*%) in metrics by excluding least confident predictions. In all three hospitals, all metrics improve except for PPV, which remains stable or decreases.

| Hospital ID | 167 |  | 199 |  | 188 |  |
| --- | --- | --- | --- | --- | --- | --- |
| DR | 100 | 83 | 100 | 94 | 100 | 90 |
| **Specificity** | 0.64 | 0.65 | 0.67 | 0.68 | 0.6 | 0.63 |
| $\Delta_{rel}\%$ | +1.6% | | +1.5% | | +5.0% | |
| **Sensitivity** | 0.6 | 0.79 | 0.7 | 0.8 | 0.88 | 0.94 |
| $\Delta_{rel}\%$ | +31.7% | | +14.3% | | +6.8% | |
| **NPV** | 0.85 | 0.94 | 0.91 | 0.95 | 0.97 | 0.99 |
| $\Delta_{rel}\%$ | +10.6% | | +4.4% | | +2.1% | |
| **PPV** | 0.33 | 0.33 | 0.33 | 0.32 | 0.28 | 0.26 |
| $\Delta_{rel}\%$ | +0.0% | | -3.0% | | -7.1% | |
| **AUC** | 0.67 | 0.79 | 0.78 | 0.83 | 0.77 | 0.85 |
| $\Delta_{rel}\%$ | +17.9% | | +6.4% | | +10.4% | |

For Hospital #167, specificity slightly improved and PPV remained stable. Other metrics peaked at DR = 83%, where they plateaued. A suggested DR of 83% would increase sensitivity (Δ*_rel_*=+31.7%), NPV (Δ*_rel_* =+10.6%) and AUC (Δ*_rel_* =+17.9%). For Hospital #199, specificity slightly increased, while sensitivity, NPV, and AUC peaked at DR = 94%. In a real-life scenario, this DR could be proposed to maximize all three metrics while maintaining a good patient proportion, improving sensitivity (Δ*_rel_* =+14.3%), NPV (Δ*_rel_*=+4.4%), AUC (Δ*_rel_* =+6.4%), and specificity (Δ*_rel_*=+1.5%), though PPV would relatively decrease by 3%.

Hospital #188 showed a similar pattern where specificity slightly increased, PPV decreased, and other metrics plateaued at DR = 90%. Figure 5c) highlights patient profiles at DR = 90% in Hospital 188, where patients with Glasgow Coma Scale (GCS) score under 7.5 and temperature over 38.1°C were excluded. Given the BaseModel’s unreliability for this profile, it would be advisable not to rely on its predictions for clinical decisions.

### 4.2 Clinical Illustration using One-Year Mortality Prediction

MED3pa supports both pre-deployment evaluation and post-deployment monitoring of a BaseModel. Here we illustrate pre-deployment use (Figure 6) with data from *Laribi et al.*^52^, where the BaseModel predicts one-year mortality upon admission. MED3pa helps refine decision-making by identifying areas of low predictive confidence.

**Figure 6:**
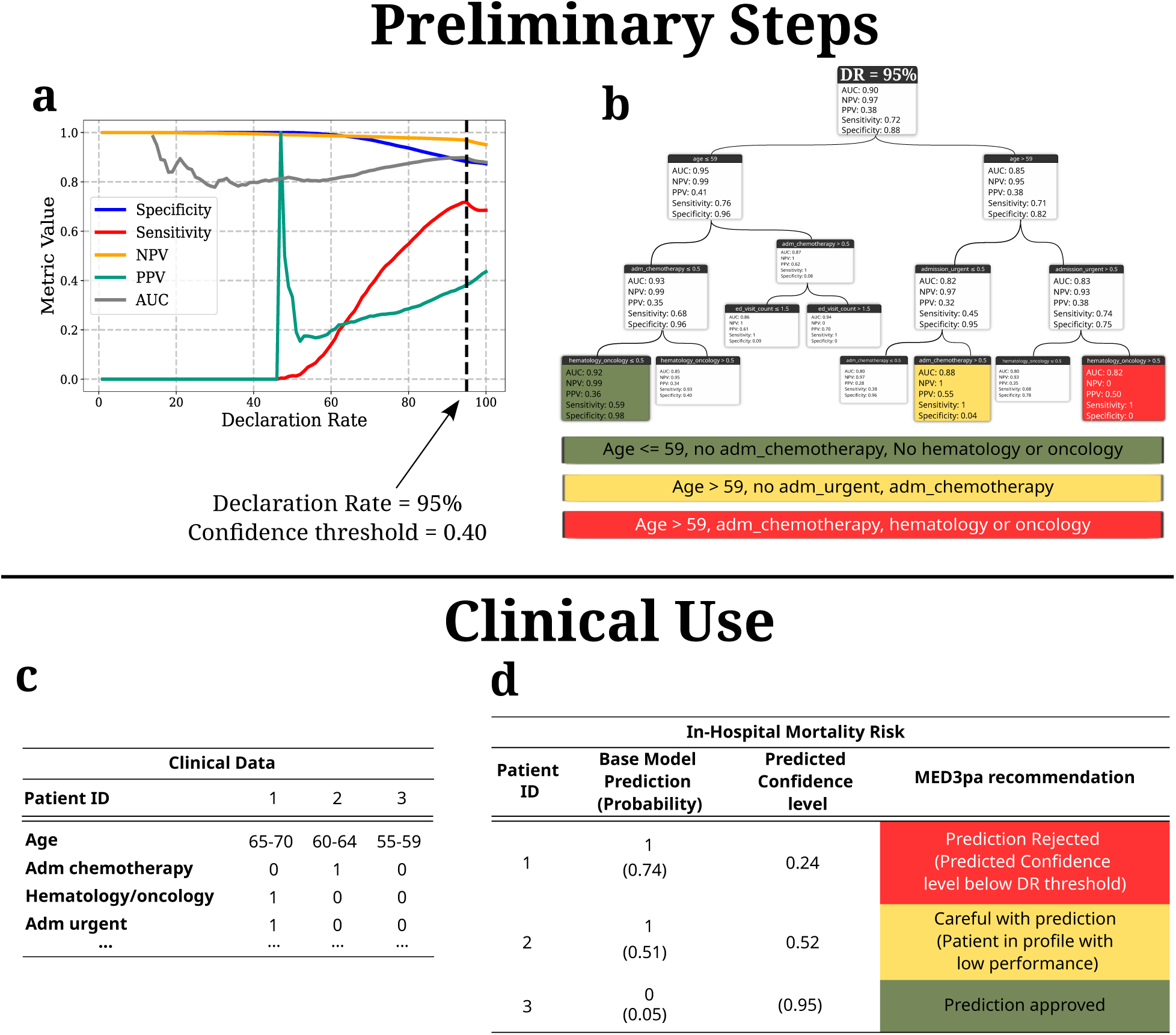
Clinical illustration of the MED3pa method for one-year mortality prediction. (a-b) Preliminary steps: This step would be done by regulatory authorities before deploying into clinical workflows. The MDR curve (a) is first used to determine an appropriate DR. A suggestion here would be DR=95%, where most metrics peak before declining, resulting in a minimum confidence threshold of 0.4. We then observe patient profiles identified by the APC model (b). The profile of patients over 59 years old, admitted urgently and with hematology/oncology diagnoses is highlighted in red due to consistently predicting mortality despite a 50% survival rate. (c-d) Example of clinical use for three representative patients examples: Once the BaseModel with the MED3pa method is deployed, the BaseModel’s predictions are to be interpreted using MED3pa recommendations. For each new patient (c), the confidence score and APC profile membership are first determined with an appropriate recommendation (d): rejection of low-confidence predictions (Patient #1), caution for patients in high-risk profiles (patient #2), and acceptance for confident predictions outside such profiles (patient #3).

Before deploying the BaseModel with MED3pa in clinical workflows, an acceptable risk threshold should be determined with regulatory authorities. This begins by analyzing overall performance and determining the optimal DR using MDR curves (Figure 6a). In this example, our goal is to improve performance without prioritizing any specific metric. Most metrics improved up to DR = 95%, after which they remained stable or declined. Based on this, we set DR at 95%.

We then examined retained patient profiles at this DR (Figure 6b). Although no profile was completely excluded, some subgroups showed significant deviations from global metrics. One such profile, highlighted in red, consisted of patients aged *>*59 years, admitted urgently with a cancer diagnosis. The BaseModel consistently predicted mortality (positive outcome) for this profile, despite an actual mortality rate of 50%, resulting in specificity and NPV of 0.

Once DR is set and high-risk profiles identified, the BaseModel with MED3pa could be deployed clinically. To illustrate practical use, we present a sample table of clinical data from three patients (Figure 6c) and show how final predictions are made (Figure 6d). For each patient, the table includes the BaseModel prediction, MED3pa’s predicted confidence, and a potential real-life recommendation.

The prediction for Patient 1 would be rejected due to low predicted confidence (below the 95% threshold). Patient 2 would receive a warning, as they belong to a high-risk profile (yellow profile in Figure 6b). Patient 3 would be approved as a reliable BaseModel prediction, as their predicted confidence exceeded the threshold and they were not part of a high-risk profile. This example shows how MED3pa highlights model limitations, supports nuanced decisions, and promotes reliable deployment.

## 5 DISCUSSION

In this study, we introduced **MED3pa**, a confidence-based method to identify and mitigate prediction errors in clinical models. Using Individualized Predictive Confidence (IPC) for patient-specific confidence, Aggregated Predictive Confidence (APC) for profile-level confidence, or their combination via Mixed Predictive Confidence (MPC), our approach helps determine when and where a BaseModel is more likely to fail during deployment.

We applied MED3pa to two classification tasks: In-Hospital Mortality (IHM) using MIMIC-IV and eICU datasets, and One-Year Mortality (OYM) using a private dataset. For both tasks, we trained a BaseModel and evaluated its reliability. Using Metrics by Declaration Rate (MDR) curves, we identified thresholds where model performance increased while excluding few patients. For example, in internal validation for IHM task, limiting predictions to the top 93% most confident cases via IPC estimates led to a 14.3% increase in sensitivity. We then used the APC model to detect patient profiles more affected by prediction errors. For instance in OYM, patients over 59 years of age, admitted urgently with cancer (Figure 6b) had 50% actual mortality rate, yet the BaseModel predicted all as positive, resulting in poor specificity and NPV. MED3pa helps uncover such model limitations unnoticed in global evaluations.

In all experiments, MED3pa improved some performance metrics while excluding only a small fraction of patients from the BaseModel’s applicability. MDR curves showed clear trade-offs between performance and patient coverage, and APC highlighted interpretable profiles showing BaseModel limitations. These findings emphasize MED3pa’s utility in exposing limitations and improving clinical safety for reliable deployment.

Our method identifies potential predictive errors and helps determine acceptable risk tolerance. Integrating MED3pa into predictive models raises awareness of limitations and may reduce systematic errors. It also supports fairness assessments by examining how confidence varies across variables such as age, sex, or ethnicity. While fairness was not the focus here, MED3pa could help detect discrimination in BaseModel predictions. The method is not limited to machine learning and could be applied to any decision-making system. It also aligns with FUTURE-AI guidelines for trustworthy, deployable AI in healthcare, including robustness, explainability, and fairness^54^.

The goal of this work is to introduce predictive confidence analysis for individual patients and profiles. While initial results are promising, several improvements remain necessary. First, the confidence metric could better account for context, such as class imbalance. Although we used a sigmoidal function (Supplementary Material E) that considers the BaseModel’s classification threshold, it is symmetrical. Incorporating asymmetry could better reflect class imbalance. The performance of MED3pa also depends on the IPC and APC models, so optimizing these components is important. While tree-based APC models produce interpretable patient profiles, they may not always reflect clinically meaningful subgroups. Exploring alternative clustering methods could help detect more granular, high-risk profiles. It would also be useful to adapt MED3pa to prioritize metrics such as sensitivity or precision, depending on clinical needs. As illustrated in Supplementary Material H, MED3pa consistently improves accuracy across all scenarios explored, but future work could adapt the method to optimize other metrics.

This is the first phase of MED3pa, our long-term goal being to better integrate all patients, not exclude them from the BaseModel’s applicability. Once we identify patients for whom the BaseModel underperforms, we aim to explain the performance gaps (e.g. covariate shifting) and refine the BaseModel accordingly. Other alternatives include comparing confidence profiles over time, or using unlabeled data to rapidly detect changes in real-time monitoring.

## 6 CONCLUSION

In this study, we introduced **MED3pa (Predictive Performance Precision Analysis in Medicine)**, a novel method that evaluates the reliability of clinical prediction models by identifying patients for whom the predictions are most reliable. MED3pa provides a structured approach to identify and characterize model confidence, improving model trustworthiness, and supporting reliable clinical decision-making. Further research will focus on explaining the sources of low confidence in model predictions, refining confidence estimation, and adapting the model to improve performance for patient profiles in which it currently under-performs.

## STATEMENTS

### Data and code availability

Software code allowing to run the experiments used to produce the results presented in this work is freely shared under the GNU General Public License v3.0 on the GitHub website at: https://github.com/MEDomics-UdeS/study_3pa. The MED3pa package developed in this study is freely shared under the GNU General Public License v3.0 at: https://pypi.org/project/MED3pa/. The data underlying this article come from both public and private sources. Publicly available data were obtained from the MIMIC-IV and eICU Collaborative Research Databases, which can be accessed upon completion of the appropriate data use agreements via PhysioNet (https://physionet.org/). The hospitalization data used for the clinical illustration via one year mortality prediction are not publicly available for confidentiality purposes overseen by the IRB (Institutional Review Board of the CIUSSS de l’Estrie—CHUS Nagano #2022-4409). However, a synthetic dataset^53^ generated using the AVATAR method^55^ in partnership with Octopize (https://www.octopize.io) is publicly shared on the Zenodo website at: https://zenodo.org/records/12954673. The publication of this synthetic dataset has been approved under project #2022-4409 with form #F2H-60510.

### Authors’ contributions

Conceptualization: OL, FCL, JFE, MV

Data curation: OL

Formal Analysis: OL

Funding acquisition: MV

Investigation: OL, LHC, LA, MV

Methodology: OL, FCL, JFE, DP, MV

Project administration: OL, FCL, JFE, DP, MV

Resources: MV

Software: OL, LHC, LA

Supervision: FCL, JFE, MV, DP

Validation: OL, DP, MV

Visualization: OL, LHC, LA

Writing – original draft: OL

Writing – review & editing: OL, MV, DP, JFE, FCL, LHC, LA

## Data Availability

All data analyzed in this study are from previously published or existing datasets, as described in the manuscript. Public datasets are available at the sources cited, while access to private datasets requires institutional approval and is not publicly available.

## Acknowledgements

The authors thank Ryeyan Taseen, MD, MSc, for data collection. We also thank Hakima Laribi, PhD student at Université de Sherbrooke, for data processing and for her helpful comments and suggestions throughout the project.

## 7 Supplementary Material

### A: Simulated Dataset description

The dataset consists of two continuous features and binary labels, chosen to facilitate visualization of the methods’ impact. For the BaseModel training data, we generated 850 data points in two classes as follows:

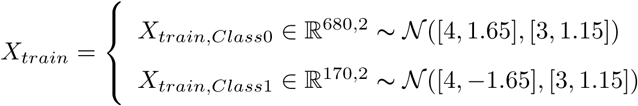

The parameters were chosen to ensure a slight overlap between the two distributions, making the classes not linearly separable. We also introduced an 80%-20% class imbalance to assess our method’s behavior in this setting.

For the test set, which simulates the deployment context of the trained **BaseModel**, we generated data following similar distributions with the same class imbalance. Additionally, we introduced an outlier cluster to the right of the main data, representing samples that do not follow the training distribution. This outlier cluster is small enough not to significantly affect overall performance metrics. The test data consists of 1000 data points generated as follows:

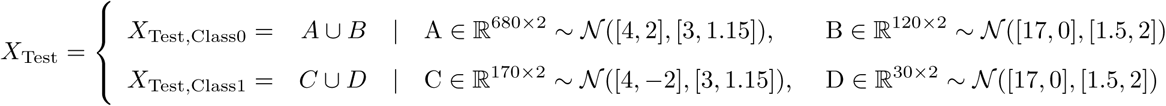

### B: Results – Empirical evaluation of confidence scores for failure detection

We evaluated whether learned confidence scores improve the detection of incorrect BaseModel predictions compared to using the BaseModel predicted score as a proxy for confidence. Predictions close to the classification threshold are typically interpreted as the most uncertain. However, relying solely on the predicted scores may be suboptimal for identifying model failures in practice. In particular, predictive scores may become miscalibrated in the deployment setting under distribution shift^56,57^, or remain well calibrated at the population level while exhibiting miscalibration within specific subgroups^58^.

The MED3pa framework addresses this limitation by estimating the risk of incorrect predictions by learning a confidence score. Specifically, MED3pa relies on three confident scores: the **Individualized Predictive Confidence (IPC)**, the **Aggregated Predictive Confidence (APC)**, and the **Mixed Predictive Confidence (MPC)**. The IPC estimates the confidence of individual predictions, the APC then aggregates these scores on a subgroup level to allow an interpretation of which patients are associated with higher risks of errors, and the MPC combines both approaches to allow a compromise between efficient and interpretable individual confidence estimation.

As a baseline, we define the confidence score derived directly from the BaseModel output. This **Base-Model confidence score** is defined as the distance between the predicted probability 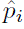 and the Base-Model’s classification threshold *τ*, normalized to ensure comparable scaling on both sides of the threshold:

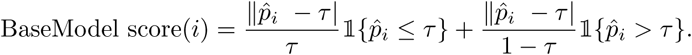

For the IPC model, we first define a target confidence score using the sigmoidal confidence function described in Supplementary Material E, which reflects the risk of incorrect predictions at the individual level. The IPC is trained as a regression model (random forest regressor) to predict this confidence score. The APC model (a regression tree) is trained to capture an aggregated version of the IPC-predicted confidence scores, learning confidence patterns across patient subgroups. Finally, the MPC confidence score is defined as the minimum between the IPC and APC predicted confidence scores.

To empirically evaluate the ability of these confidence scores to detect incorrect predictions of the Base-Model, we conducted experiments across all datasets used in this study. For each dataset, the procedure was repeated 100 times. In each repetition, the evaluation set was randomly split in half. One half was used to train the IPC and APC models, while the remaining half was used to evaluate the discrimination performance of all confidence scores between correct and incorrect predictions. The performance were compared using the Area Under the Receiver Operating Characteristic Curve (AUC), with higher AUC values indicating a better ability to identify predictions at higher risk of failure.

The reported results correspond to the mean AUC and standard deviation across the 100 repetitions and are summarized in Table 1. Across all datasets, the confidence predicted by IPC, APC, and MPC consistently outperformed the BaseModel confidence score. IPC achieves the highest discrimination performance, while APC still provides useful confidence estimates by aggregating confidence patterns. The MPC offers an interesting compromise between the IPC and APC, maintaining higher discrimination abilities while incorporating aggregated information.

**Table 1:**
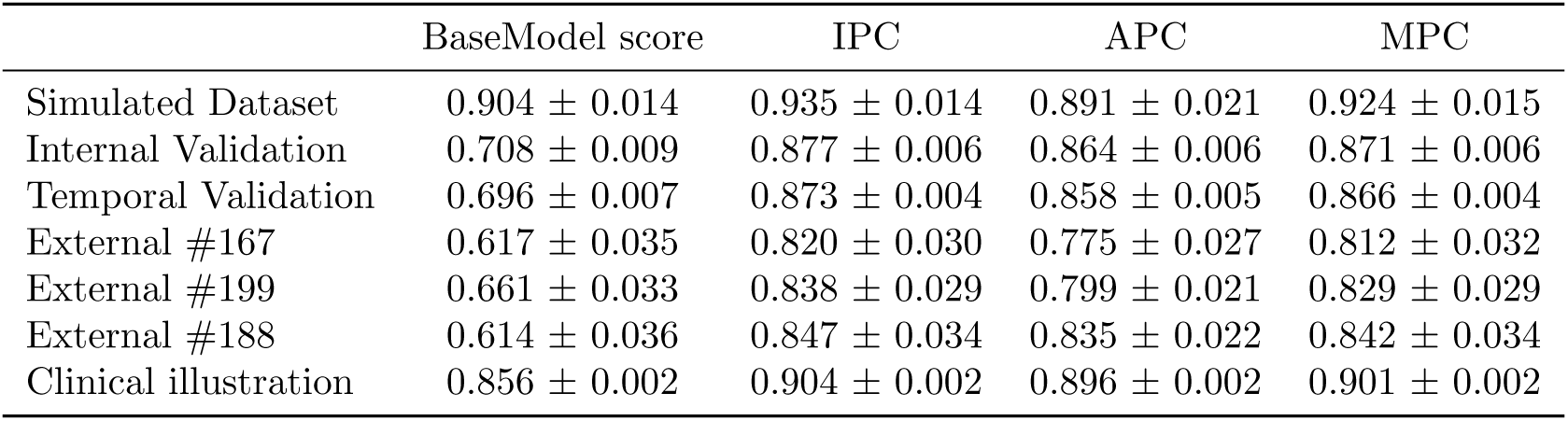
AUC values for detecting BaseModel prediction errors using the BaseModel predicted score, as well as the IPC-predicted, APC-predicted and MPC-predicted confidence across all datasets used in this study. The MPC-predicted confidence is defined as the minimum between IPC-predicted confidence and APC-predicted confidence. (mean ± standard deviation over 100 random half-sample splits).

### C: Predictive confidence models training

The training procedure for the **Individualized Predictive Confidence** (IPC), **Aggregated Predictive Confidence** (APC) and the **Mixed Predictive Confidence** (MPC) models is designed to suit the intended application of the method. This section provides training strategies for the MED3pa method, whether the goal is to predict individual-level confidence, profile-level confidence, or a compromise between those two.

**Individual-Level predictions only** When the goal is to make confidence predictions for individual patients, we train the IPC on a subset of the evaluation dataset. Specifically, we use a fraction (we suggest using half) of the available evaluation dataset to train the IPC model. The remaining data is used to compute Metrics by Declaration Rate (MDR) curves to evaluate the BaseModel’s performance for different levels of confidence. This approach ensures that the IPC model can be used to predict the confidence of individual predictions, while reducing the risk of overfitting the data in the results of MDR curves.

**Profile-Level predictions only** If the goal is to predict confidence values for profiles of patients (sub-groups), similarly to the previous scenario, we train the APC on a fraction (we also suggest using half) of the available evaluation dataset. The remaining dataset is then used to compute the MDR curves, showing the impact of excluding whole profiles on the performance of the BaseModel.

**Compromise between Individual and Profile-Level predictions** For an approach that incorporates both individual-level and profile-level confidence predictions, we first train the IPC model on a first fraction (we suggest using half) of the evaluation dataset to capture individual-level confidence. We then use the same data to train the APC model. The APC is however trained to predict the IPC-derived confidence. This ensures consistency between the profiles created by the APC model and the predictions of the IPC. We then combine the IPC and APC models following the desired approach for the MPC (minimum confidence, averaging, …). The best strategy can be selected either by evaluating which combination yields the most reliable confidence estimates, or choosing the one that aligns best with the clinical context. The MDR curves are then computed using the confidence predictions of the Mixed Predictive Confidence model with remaining subset of data.

### D: Algorithmic details

This section provides additional details on the experimental setup and algorithmic components used in each MED3pa evaluation. Supplementary Figure 1 illustrates the step-by-step application of the MED3pa method, starting from a pretrained BaseModel to the generation of individualized and aggregated predictive confidence estimates. Supplementary Table 2 complements this figure by specifying the exact models and parameters used across all four evaluation settings: internal validation (IV), temporal validation (TV), external validation (EV), and the clinical illustration.

**Figure 1:**
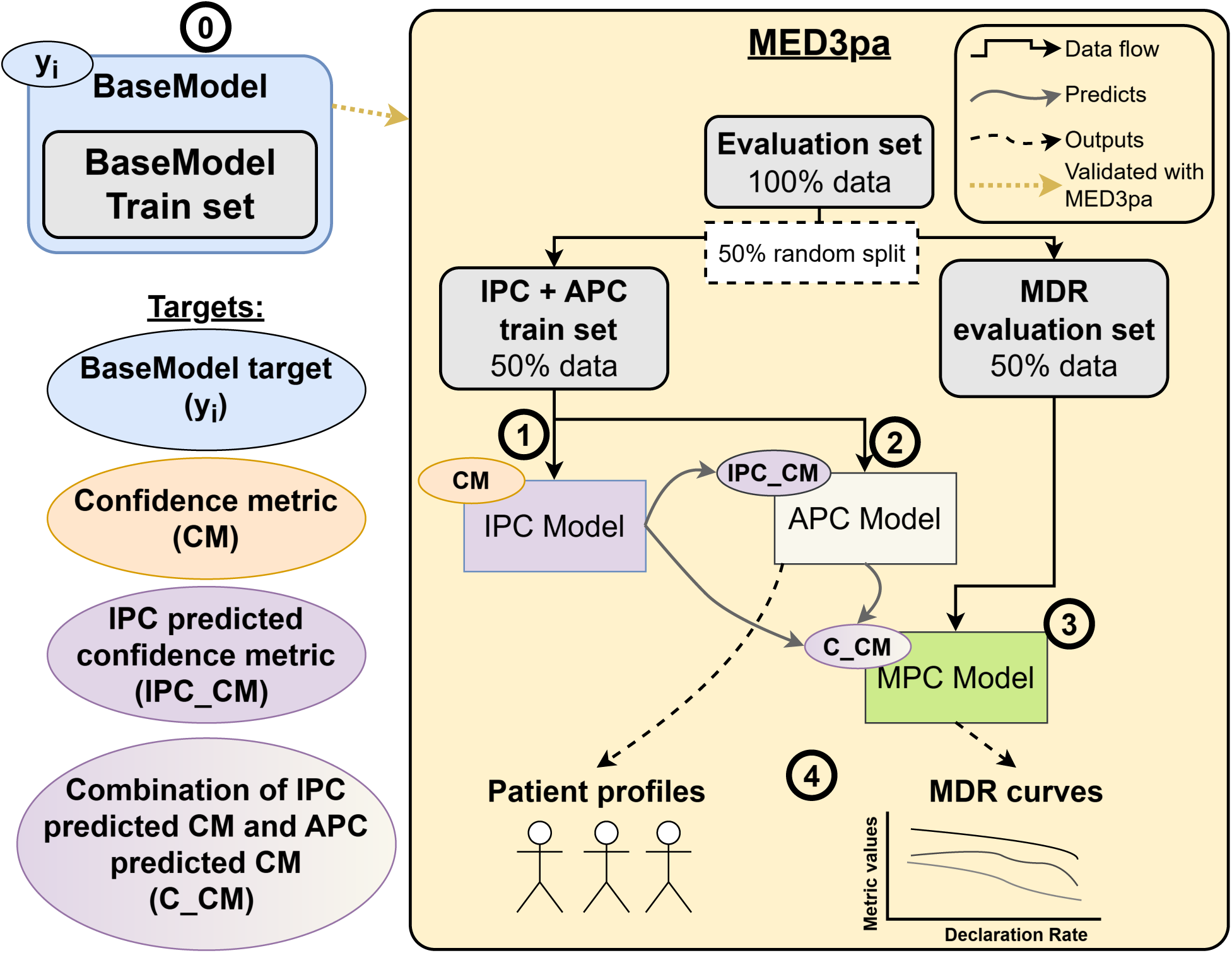
Experimental setup of the MED3pa method used in each evaluation. A pretrained BaseModel is first trained on its respective training set to predict the target variable *y_i_* (Step 0). The MED3pa method is then applied to assess the BaseModel’s reliability. The evaluation set is randomly split in two: one half is used to train the Individualized Predictive Confidence (IPC) model (Step 1), which learns to estimate a predictive confidence metric (CM) for each patient. The APC model (step 2) is trained on the same subset to predict IPC-derived confidence as the subgroup level. The Mixed Predictive Confidence (MPC) strategy (Step 3) combines IPC and APC outputs (C CM). Finally, MDR curves as well as profile-specific metrics are computed using the remaining half of the dataset (step 4).

**Table 2:** Experimental setup details for each MED3pa evaluation setting. Sigmoidal function refers to the sigmoidal confidence function provided in Supplementary Material E. IV: Internal Validation; TV: Temporal Validation; EV: External Validation; XGBoost: eXtreme Gradient Boosting; RFC: Random Forest Classifier; RFR: Random Forest Regressor; RT: Regression Tree.

| **Component** | **IV** | **TV** | **EV** | **Clinical Illustration** |
| --- | --- | --- | --- | --- |
| **Confidence Metric** | Sigmoidal function | Sigmoidal function | Sigmoidal function | Sigmoidal function |
| **BaseModel** | XGBoost | XGBoost (from IV) | XGBoost (from IV) | RFC |
| **IPC Model** | RFR | RFR | RFR | RFR |
| **APC Model** | RT | RT | RT | RT |
| **MPC Strategy** | Min(IPC, APC) | Min(IPC, APC) | Min(IPC, APC) | Min(IPC, APC) |

### E: Sigmoidal confidence function

We use a sigmoidal function to transform model error indicators into a continuous confidence score between 0 and 1. This smooth mapping allows for a flexible assessment of prediction confidence while incorporating the BaseModel classification threshold, which is central to our MED3pa framework. We define the sigmoidal confidence function as

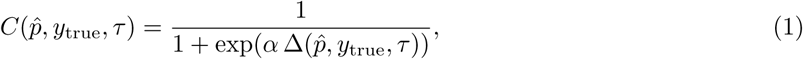

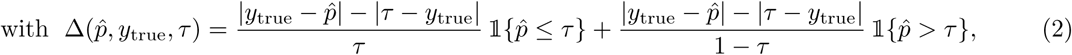

where *α* is the slope of the function, *y_true_* is the ground truth label, 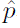 is the BaseModel’s probability estimate, and *τ* represents the classification boundary of the BaseModel. We defined this confidence function to incorporate the classification threshold of the BaseModel to ensure that the confidence measure aligns with the actual decision rule used by the BaseModel. The slope parameter was set to *α* = 5 ln(3) so that when the predicted probability deviates by 20% of the classification threshold, the confidence value is 0.75 when the binarized prediction matches the ground truth, and 0.25 otherwise.

### F: In-hospital mortality dataset description

For the in-hospital mortality prediction task, we used MIMIC-IV^48^ and eICU-CRD^49^ datasets. We used the same predictors as used in the Simplified Acute Physiology Score (SAPS) II score^47^. A detailed descriptive analysis of these variables and the SAPS II encoding applied to each variable is provided in Supplementary Table 3. We then applied the same exclusion criteria as *Zeng et al.*^50^, which are shown in Supplementary Figure 2. First, we selected only the first ICU stay for patients with a single hospitalization. We then excluded patients who did not meet the sepsis-3 criteria^59^. Finally, we only included patients aged 16 to 89 years, having an ICU stay duration of at least 24 hours and having at least 30% of predictor values present. Missing data were imputed using a K-nearest neighbors (K-NN) algorithm.

Although the precise admission years for patients in the MIMIC dataset are not available, admissions are categorized into four distinct anchor year groups, allowing us to differentiate between cohorts based on their respective admission years. As shown in Supplementary Figure 2, we used the first two cohorts (2008-2010 and 2011-2013) to train and validate a BaseModel classifier. Admissions in these cohorts were randomly split in half, using 50% of them to train the BaseModel, while assessing the BaseModel’s **internal validity** using the other half with the MED3pa method. The latter two cohorts (2014-2016 and 2017-2019) served as a temporal test set, thereby assessing the BaseModel’s **temporal validity** with the MED3pa Method. For the eICU-CRD dataset, we group patients by their respective hospitals of admission, allowing us to examine the external validity of the BaseModel trained on MIMIC in the context of each individual hospital. We evaluate **external validity** using only hospitals with at least 225 hospitalizations after processing steps.

**Table 3:** Descriptive analysis of the SAPS II^47^ variables used for the in-hospital mortality task. Results are shown for all three evaluation sets used with the MED3pa method: internal validation (IV), temporal validation (TV), and external validation (EV). We report the mean and standard deviation for each continuous variable, and the proportion of patients for categorical and binarized variables. We also show the SAPS II^47^ encoding applied to each variable according to the SAPS-II^47^ score. GCS: Glasgow Coma Scale. BUN: Blood Urea Nitrogen. WBC: White Blood Cell count. AIDS: Acquired immunodeficiency syndrome.

| Variable | MIMIC-IV |  | eICU-CRD | SAPS II<sup>47</sup><br>encoding | Retained value |
| --- | --- | --- | --- | --- | --- |
|  | IV<br>(n=4,476) | TV<br>(n=7,319) | EV<br>(n=2,583) |  |  |
| Age | 65.4 ± 15.5 | 64.1 ± 15.7 | 62.7 ± 15.8 | – | Age |
| Heart Rate (bpm) | 91.5 ± 32.5 | 91.7 ± 33.2 | 105.5 ± 33.1 | <70 | Lowest value |
|  |  |  |  | ≥ 70 | Highest value |
| Systolic BP (mmHg) | 99.2 ± 35.2 | 98.5 ± 33.9 | 93.1 ± 33.6 | <100 | Lowest value |
|  |  |  |  | ≥100 | Highest value |
| Temperature (°C) | 37.6 ± 0.8 | 37.5 ± 0.8 | 37.8 ± 0.9 | – | Highest value |
| GCS | 13.1 ± 3.4 | 13.4 ± 3.0 | 13.0 ± 3.1 | – | Lowest value |
| PaO<sub>2</sub>/FiO<sub>2</sub> (mm Hg/%) | 3.7 ± 4.1 | 4.1 ± 4.3 | 1.2 ± 3.1 | <100 | 11 |
|  |  |  |  | ≥100 & <200 | 9 |
|  |  |  |  | ≥200 | 6 |
|  |  |  |  | Not ventilated | 0 |
| Urine Output (mL/day) | 1906.8 ± 1241.5 | 1822.1 ± 1332.8 | 1591.8 ± 1303.9 | – | Total Urine |
| BUN (mg/dl) | 27.9 ± 22.3 | 28.3 ± 23.4 | 37.6 ± 27.9 | – | Highest value |
| WBC (x 10<sup>3</sup>mm<sup>3</sup>) | 12.5 ± 11.0 | 14.0 ± 12.9 | 15.8 ± 12.4 | ≥20.0 | Highest value |
|  |  |  |  | <20.0 | Lowest value |
| Sodium (mEq/L) | 137.9 ± 5.8 | 137.7 ± 6.5 | 136.4 ± 7.3 | ≥145 | Highest value |
|  |  |  |  | <145 | Lowest value |
| Potassium (mEq/L) | 4.4 ± 0.8 | 4.5 ± 0.8 | 4.4 ± 1.0 | <3.0 | Lowest value |
|  |  |  |  | ≥3.0 | Highest value |
| Bicarbonate | 21.9 ± 4.6 | 20.8 ± 4.7 | 20.0 ± 5.7 | – | Lowest value |
| Bilirubin | 2.5 ± 3.7 | 2.3 ± 4.1 | 1.7 ± 3.0 | – | Highest value |
| AIDS | 0.8% | 0.5% | 0.4% | Presence | Binarized |
| Metastatic Cancer | 6.8% | 6.0% | 1.4% | Presence | Binarized |
| Hematologic Malignancy | 3.9% | 3.0% | 2.4% | Presence | Binarized |
| Admission Type | 5.0% | 4.2% | 1.9% | Scheduled surgical | 0 |
|  | 64.6% | 66.8% | 96.4% | Medical | 6 |
|  | 30.3% | 29.0% | 1.7% | Unscheduled surgical | 8 |
| In-hospital mortality | 17.4% | 18.8% | 20.3% |  |  |

**Figure 2:**
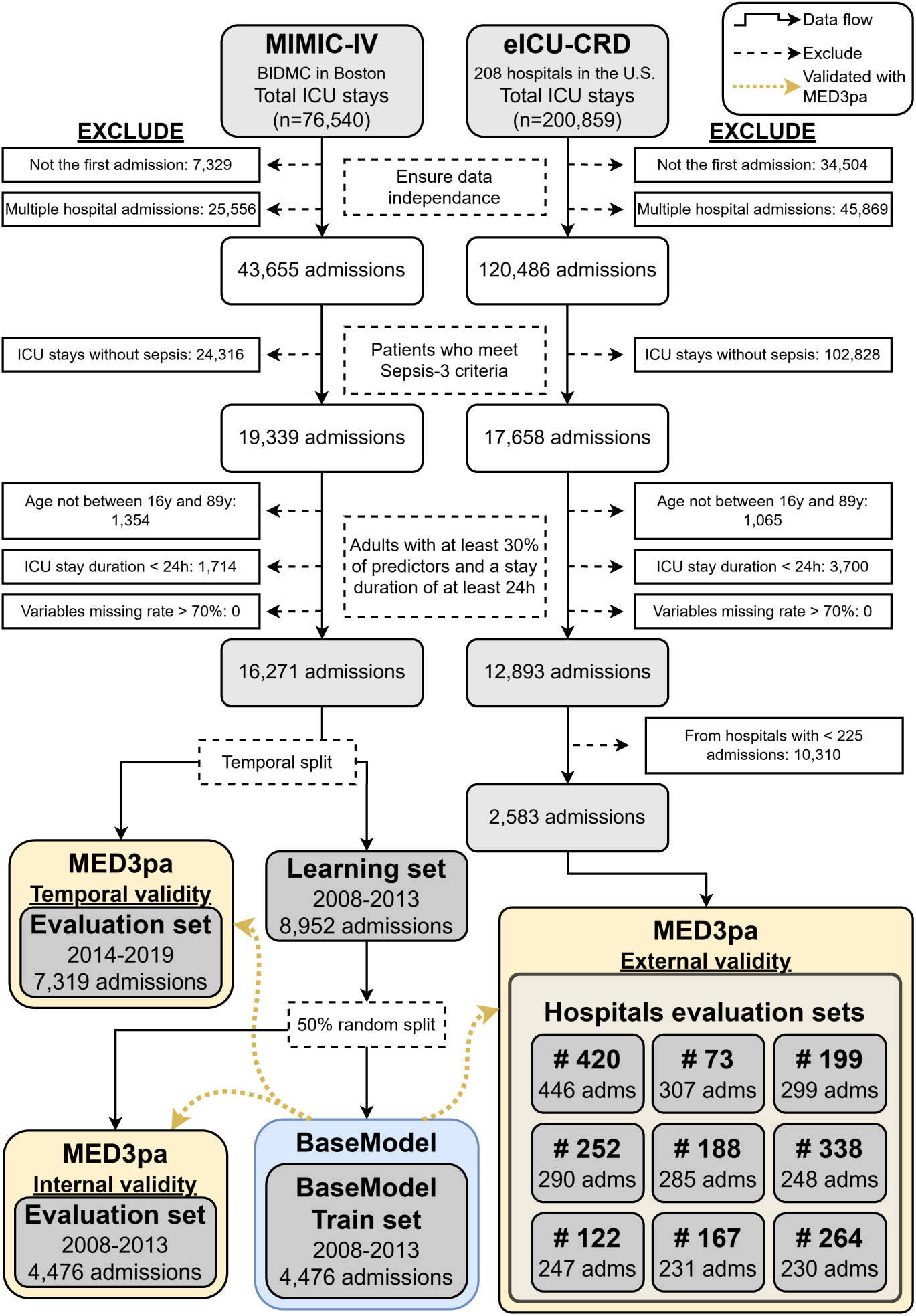
Data processing of MIMIC-IV and eICU-CRD datasets for the in-hospital mortality classification task. We first excluded admissions following *Zeng et al.*^50^. We then divided the remaining data from MIMIC-IV into two temporal cohorts: admissions from 2008–2013 were used as the learning set, while admissions from 2014–2019 formed the temporal evaluation set. The learning set was further split randomly into two equal parts: one half was used to train the BaseModel, and the other half was reserved for internal validation using the MED3pa method. For external validation, data from eICU-CRD were grouped by hospital. Only hospitals with at least 225 admissions were retained, and the selected hospitals were used to assess external model performance.

### G: One-year mortality dataset description

For the one-year mortality classification task, we used the data from *Laribi et al.*^52^, which comprises EHRs and administrative data from an integrated university hospital network in Sherbrooke, Quebec, Canada. Supplementary Table 4 provides a detailed descriptive analysis across the full dataset of the demographic and admission characteristic features, as well as four major comorbidities, taken from *Laribi et al.*^52^. Supplementary Figure 3 shows the processing applied to the data, as well as to which data was used to train the BaseModel, and which data was used as the evaluation set for the temporal validation of the BaseModel using the MED3pa method.

**Table 4:** Taken from *Laribi et al.*^52^. Descriptive analysis of the demographics and admission characteristics features along with four major comorbidities on the full dataset. We present the mode of each categorical feature along with its proportion in the dataset, and the mean of each continuous feature along with its standard deviation. The *p*-values are computed using the Welch’s t-test^60^ for continuous features (age, ambulance admissions count, ED visits count, weeks recently hospitalized) and the Pearson’s chi-squared test^61^ for categorical and binary features with the scipy^62^ Python library.

| Variable | All<br>(n=250,812) | Survivors<br>(n=214,095) | Deceased<br>(n=36,717) | *p*-value |
| --- | --- | --- | --- | --- |
| Demographics |  |  |  |  |
| Age | 61.12 ± 20.07 | 58.98 ± 20.18 | 73.62 ± 13.93 | < 0.001 |
| Sex | Female (54 %) | Female (55 %) | Male (54 %) | < 0.001 |
| Admission characteristics |  |  |  |  |
| Ambulance admission | 0 (71 %) | 0 (74 %) | 1 (51 %) | < 0.001 |
| Flu season | 0 (75 %) | 0 (75 %) | 0 (74 %) | < 0.001 |
| ICU admission | 0 (97 %) | 0 (97 %) | 0 (95 %) | < 0.001 |
| Urgent 30-d readmission | 0 (90 %) | 0 (92 %) | 0 (79 %) | < 0.001 |
| Ambulance admissions count | 0.23 ± 0.75 | 0.17 ± 0.64 | 0.56 ± 1.19 | < 0.001 |
| ED visits count | 0.8 ± 1.52 | 0.7 ± 1.42 | 1.38 ± 1.89 | < 0.001 |
| Weeks recently hospitalized | 0.29 ± 0.99 | 0.21 ± 0.85 | 0.71 ± 1.5 | < 0.001 |
| Living status | Home (48 %) | Unknown (50 %) | Home (59 %) | < 0.001 |
| Admission service | Cardiology (13 %) | Obstetrics (15 %) | I.M. (14 %) | < 0.001 |
| Admission type | Urgent (65 %) | Urgent (60 %) | Urgent (89 %) | < 0.001 |
| Major comorbidities |  |  |  |  |
| Dementia | 0 (97 %) | 0 (98 %) | 0 (93 %) | < 0.001 |
| Congestive heart failure | 0 (94 %) | 0 (95 %) | 0 (86 %) | < 0.001 |
| Metastatic solid cancer | 0 (98 %) | 0 (99 %) | 0 (91 %) | < 0.001 |
| Asthma | 0 (97 %) | 0 (97 %) | 0 (96 %) | < 0.001 |

I.M.: Internal Medicine.

**Figure 3:**
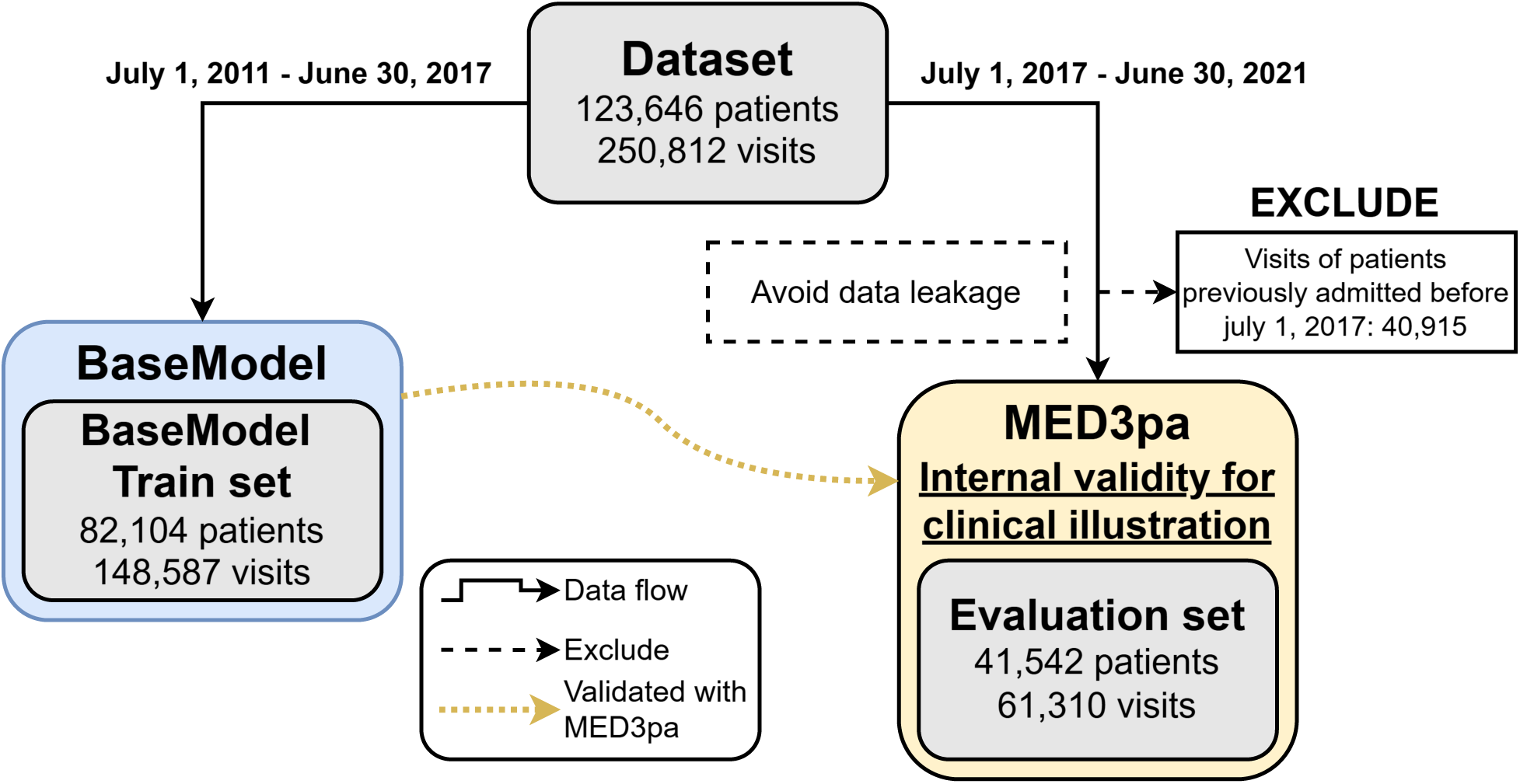
Data processing of the data from *Laribi et al.*^52^ for the one-year mortality classification task. We first temporally divide the dataset into a BaseModel train set and an evaluation set for the temporal validation of the BaseModel with the MED3pa method. We then excluded patients in the evaluation set that were previously admitted in the BaseModel train set to avoid data leakage. We finally used the trained BaseModel on the evaluation set to validate its reliability for the clinical illustration.

### H: Accuracy by Declaration Rate

This figure combines results from all experiments in the study, illustrating that accuracy globally increases as predictions with the lowest predicted confidence are progressively excluded.

**Figure 4:**
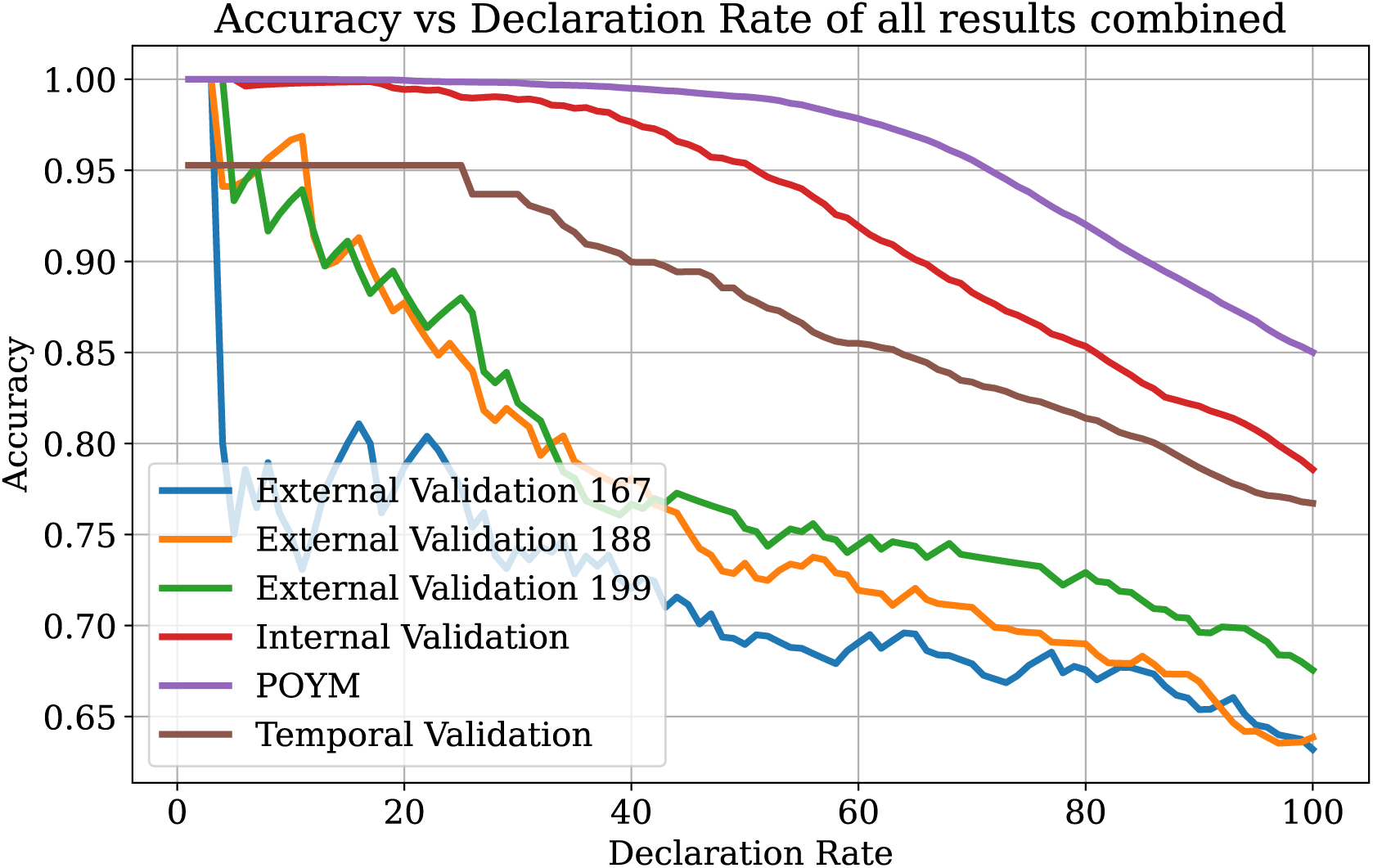
Accuracy by Declaration Rate of all experiments.

